# Integrated Bite Case Management within a One Health Framework for Rabies Elimination at the Primary Care Level in Kerala, India: An Implementation Research Protocol Using a Stepped-Wedge Cluster Randomized Design

**DOI:** 10.64898/2026.01.08.26343709

**Authors:** Zinia Thajudeen Nujum, Martha Luka, Dhanya Raghunathan, Mufeeda Beegum, S Harikumar, Prapti Bajaj, Balaji Chandrashekar, K Rajamohanan, Thomas Mathew, Biju Soman, Miqdad Asaria, K. Rekha, Andrew David Gibson, M.R. Sreenath, Arun Jose, Rony Ray John, S Aparna, P.R. Prathiush, Swapna Susan Abraham, S.B. Narayanan, Ummuselma Chungath, P S Indu, U Anuja, S Remadevi, K Vijayakumar, S.S. Aishwarya, Katie Hampson

## Abstract

**Background:** Rabies remains a persistent public health threat in India, while gaps in surveillance and coordination of control measures undermine progress towards ending human rabies deaths. The World Health Organization advocates a One Health approach, combining mass dog vaccination, post-exposure prophylaxis (PEP), and Integrated Bite Case Management (IBCM). IBCM is not yet incorporated into India’s national rabies action plan for rabies elimination (NAPRE). This study seeks to generate robust evidence on the effectiveness, cost-effectiveness, feasibility, facilitators and barriers of IBCM implemented within Kerala’s Rabies Control Program.

**Methods:** This will be an implementation-research, adopting a stepped-wedge cluster randomized controlled trial (RCT) design. IBCM will be implemented across six administrative blocks in Thiruvananthapuram District, Kerala, across diverse rural and urban settings, and including tribal population representation. The IBCM intervention includes stakeholder training and support, and active animal surveillance aligned with WHO guidance. Suspect or probable rabies exposures presenting at selected health facilities and identified via a community-based hotline will trigger investigations and responses including dog vaccination and sensitization. Prior to IBCM implementation, baseline data will be collected on management of dog bites, bite patient incidence and health case seeking behaviours. Hospital-based event tracking of bite cases will be conducted at enrolled public health facilities before and after IBCM implementation. The resulting RCT data will be used to model the impacts and cost-effectiveness of IBCM, including the potential for cost savings through judicious PEP, if scaled up across Kerala. To understand the barriers and facilitators to IBCM implementation, we will use in-depth interviews and focus group discussions with stakeholders.

**Discussion:** There is an urgent need to strengthen rabies surveillance, including coordination across health and animal health sectors and outbreak responses to accelerate rabies elimination. IBCM is expected to lead to improved PEP completion, increased detection and laboratory confirmation of rabid dogs, strengthened dog vaccination coverage, and targeted outbreak responses in high-risk areas. This RCT will provide critical evidence to inform policy, support scale-up, generate insights into contextual barriers and facilitators for IBCM implementation, and guide India’s One Health rabies elimination strategy. Unlike the Goa model that relies on hotline reporting, this study evaluates a health-sector–initiated IBCM approach that identifies cases through dog-bite patients presenting to health facilities, enabling more comprehensive One Health integration and greater event capture.

**Trial registration:** CTRI registration No. REF/2024/04/081804

## Introduction

Rabies is a neglected tropical disease endemic in over 150 countries, causing an estimated 59,000 deaths annually and an economic burden of around USD 8.6 billion.[1] Current rabies prevention strategies largely rely on post-exposure prophylaxis (PEP), which involves vaccinating individuals following exposure to potentially rabid animals. The One Health approach is a unifying approach that aims to sustainably balance and optimise the health of humans, animals and ecosystems and emphasizes integrated interventions across human, animal, and environmental sectors.[2,3] Mass dog vaccination, with a target coverage of 70% is needed to eliminate dog-mediated rabies,[4] which accounts for 99% of human rabies deaths.[1] However, global investment in dog vaccination remains disproportionately low^1^, while spending on PEP remains high.

Coordinated surveillance between human and animal health sectors is crucial for detection of animal rabies cases and for evaluating the effectiveness of control programmes. This forms the basis of ‘Integrated Bite Case Management’ (IBCM), an advanced One Health surveillance method that connects health professionals, veterinary professionals and the community.[5–10]Modelling work suggests that a combination of mass dog vaccination, improved PEP access and IBCM is the most cost-effective strategy for reducing human rabies deaths.[11] The World Health Organization (WHO) has underscored the importance of IBCM in its One Health open course module.[12] However, for successful inclusion of IBCM into national rabies control and prevention programs, there is a need for demonstration projects supported by implementation research involving key stakeholders such as program managers, political leaders and intersectoral officials.

India bears a disproportionate burden of global rabies deaths. However, the latest countrywide survey suggests an overall decline to around 6000 rabies deaths per year. [14] This is attributable to improvements in public health infrastructure, awareness, human rabies vaccine availability, and intradermal vaccination methods in many parts of the country. To further this progress, India has established the National Action Plan for Rabies Elimination (NAPRE), aiming to eliminate dog-mediated rabies deaths by 2030. While NAPRE focuses on mass dog vaccination and PEP provisioning, it does not explicitly incorporate IBCM as part of its surveillance strategy.[15]

Kerala, India’s most advanced state in terms of Human Development Index, reports around 1.6% of its population sustaining animal bites annually according to the Integrated Disease Surveillance Program (IDSP) data for 2018-2022 (pers. comm, Department of Public Health, Directorate of Health Services). Annual rabies deaths in Kerala range between 5 and 26, since 2018.[16] The state’s robust primary healthcare system, decentralized governance, and active community participation create an ideal environment for implementing One Health strategies like IBCM. As part of the Rebuild Kerala Initiative, a World Bank funded project to support socioeconomic development in Kerala, the government launched a Centre for One Health.[17] Nearly 900,000 people in Kerala receive PEP every year (pers comm, Department of Public Health, Directorate of Health Services), placing a growing financial burden on the government. In this context, introducing IBCM offers an opportunity to improve the efficiency of PEP use by targeting it more appropriately, while strengthening the link between human and animal health surveillance and improving dog vaccination to address the source of transmission. This integrated One Health approach could make rabies prevention efforts more effective, sustainable, and aligned with the state’s One Health vision.

A recent systematic review on the cost-effectiveness of One Health rabies interventions highlighted a scarcity of high-quality implementation studies on rabies.[18] This study is designed to fill that evidence gap, by carrying out a stepped-wedge cluster randomized controlled trial (RCT) of IBCM implementation in Kerala. It aims to examine the effectiveness of IBCM for improving rabies prevention, control and surveillance, in terms of PEP completion among bite victims, enhanced dog vaccination and detection of animal rabies cases. We further aim for a cost-effectiveness modelling and identification of the facilitators and barriers influencing the implementation of IBCM, via implementation outcomes of acceptability, adoption, appropriateness, feasibility, fidelity, penetration, and sustainability.

## Methods

### Study Design

We will use a pragmatic, stepped-wedge cluster RCT to evaluate the implementation of IBCM in Kerala, India. The study will involve six clusters and will span a pre-implementation period(Preparatory phase) prior to the stepped rollout of IBCM (Implementation Phase) followed by a period of sustained implementation (Sustenance Phase); (Fig 1). Data collection will continue throughout. Baseline data collected prior to IBCM implementation will capture existing practices in the management of dog bite cases in the absence of IBCM and a cross-sectional survey (S1) will assess the incidence of animal bites and related health care-seeking behaviour to inform analyses. The sequential rollout of IBCM across clusters will be determined through randomized allocation. An embedded process evaluation will explore the acceptability, adoption, appropriateness, feasibility, fidelity, penetration, and sustainability of IBCM implementation guided by the RE-AIM framework.[19]. The measured outcomes will be further used to model the impacts and cost-effectiveness of IBCM if scaled up across the state of Kerala.

**Fig 1.**
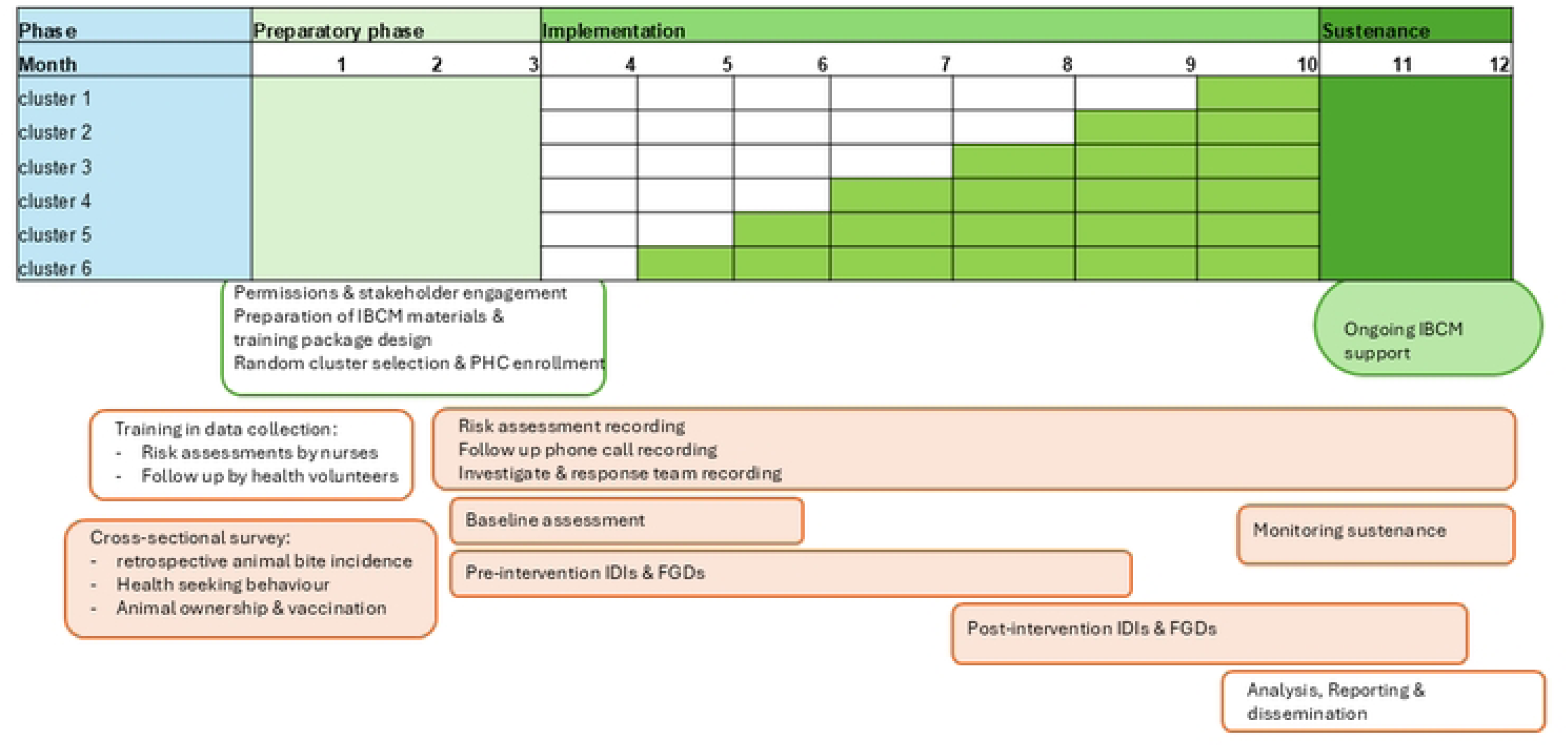
Study design for stepped-wedge cluster randomized controlled trial of Integrated Bite Case Management in Kerala, India.

### Study setting and population

The study will be conducted in Thiruvananthapuram district, Kerala, India (Fig 2). Kerala comprises 14 districts, among which Thiruvananthapuram consistently reports the highest burden of dog bites. State-level data from the IDSP shows an annual incidence of 15 animal bite patients and 8.3 dog bite patients seeking hospital care per 1,000 population in Thiruvananthapuram, almost all of whom receive PEP (pers. comm, Department of Public Health, Directorate of Health Services).

**Fig 2.**
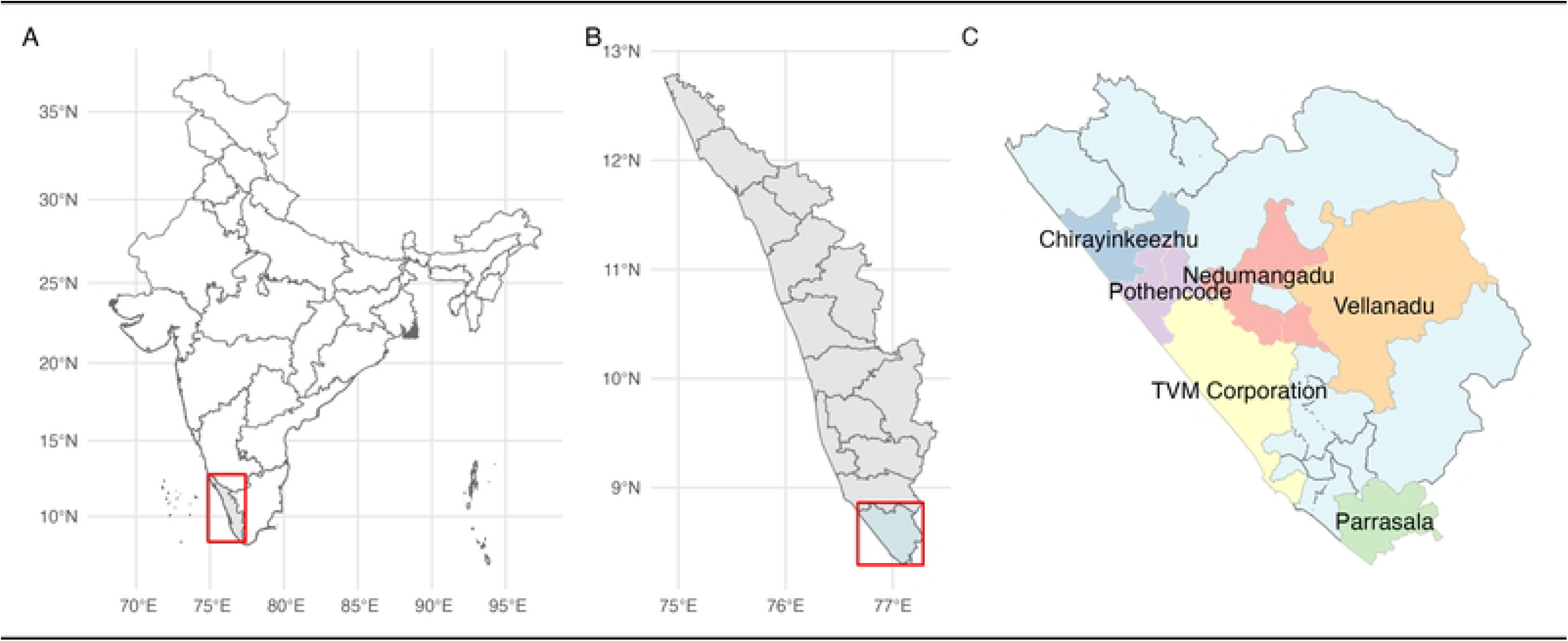
Map of the study setting: **(A)** India showing Kerala; **(B)** Kerala showing Thiruvananthapuram district; and **(C)** Thiruvananthapuram district showing the **six clusters** selected for Integrated Bite Case Management (IBCM) implementation, Kerala, India.

Primary health care in the district is delivered through Primary Health Centres (PHCs),[20] some of which have been upgraded to Family Health Centres (FHCs).[21] Typically, five to six PHCs/FHCs are grouped under a Block Primary Health Centre or a Community Health Centre (CHC).[20] Thiruvananthapuram district comprises 11 rural blocks and three urban administrative zones.[22] We will select six clusters (Table 1), with each defined as an administrative block/area (a sub-district unit of governance in India). These will be stratified to include five rural blocks, including one with a sizable tribal population purposively selected to ensure adequate representation of tribal communities, and one urban administrative zone (Thiruvananthapuram corporation).

**Table 1.** Scoring to Select Study Sites Based on Animal and Human Rabies Data and Dog Bite Cases.

| Sl. No. | Site | Lab confirmed Animal Rabies reported (2022) | Lab confirmed Animal Rabies reported (2023) | Human Rabies Deaths 2022&23 | No of times the area figured in the top 10 areas based on Dog Bite Cases/Month (Jan–Aug 2022) | Total Score* | Rank |
| --- | --- | --- | --- | --- | --- | --- | --- |
| 1 | Parassala |  | 2 | 1 | 3 | 6 | 4 |
| 2 | Perumkadavila | 1 |  |  | 1 | 2 |  |
| 3 | Athiyannoor |  | 1 |  | 1 | 2 |  |
| 4 | Nemom |  |  |  | 0 | 0 |  |
| 5 | Pothencode | 3 |  | 1 | 4 | 8 | 3 |
| 6 | Vellnad** |  |  |  | 0 | 0 |  |
| 7 | Nedumangad | 1 | 3 | 1 | 5 | 10 | 2 |
| 8 | Vamanapuram |  | 1 | 1 | 2 | 4 |  |
| 9 | Kilimanoor |  | 1 | 1 | 2 | 4 |  |
| 10 | Chirayinkeezhu | 3 |  |  | 3 | 6 | 5 |
| 11 | Varkala | 1 |  | 1 | 2 | 4 |  |
| 12 | Thiruvananthapuram Corporation | 1 | 5 | 3 | 9 | 18 | 1 |
| 13 | Nedumangad Municipality | 2 |  |  | 2 | 4 |  |
| 14 | Attingal Municipality |  | 1 | 1 | 2 | 4 |  |

\*Using data from three sources namely lab-confirmed animal rabies cases in 2022 and 23 (from State Institute of Animal Diseases), Human cases reported in 2022 and 2023 and reported dog bite cases/month reported to veterinary hospitals/dispensaries (January to August 2022 – if the area was listed in the based in top 10 areas according to dog bite cases and if the numbers were above 15 cases/month it was given as score of 1 , each month that it appeared considered) a scoring system was developed. Each time a block was represented in the data a score of 1 was given.
\*\* purposively included sixth cluster for tribal representation

The study participants will be the patients presenting to PHCs with dog bite(s)/ scratch(es)/ other potentially rabies-risk contacts with dogs (defined as a lick to abraded skin or mucosal membrane or substantive contact with a dog identified as rabid) and administered PEP. Exposures from other animals will be excluded. For pragmatic purposes, the first two dog bite patients presenting each day to the enrolled PHCs will be recruited to the IBCM intervention.

For the process evaluation study participants will include purposively selected stakeholders involved in IBCM (Fig 3) including political leaders at state, district and local levels, program managers from the human and animal health sectors, medical officers and health workers from both the Directorate of Health Services (DHS) and Animal Husbandry Department (AHD), Accredited Social Health Activists (ASHA) workers and individuals exposed to animal bites for beneficiary case studies.

**Fig 3.**
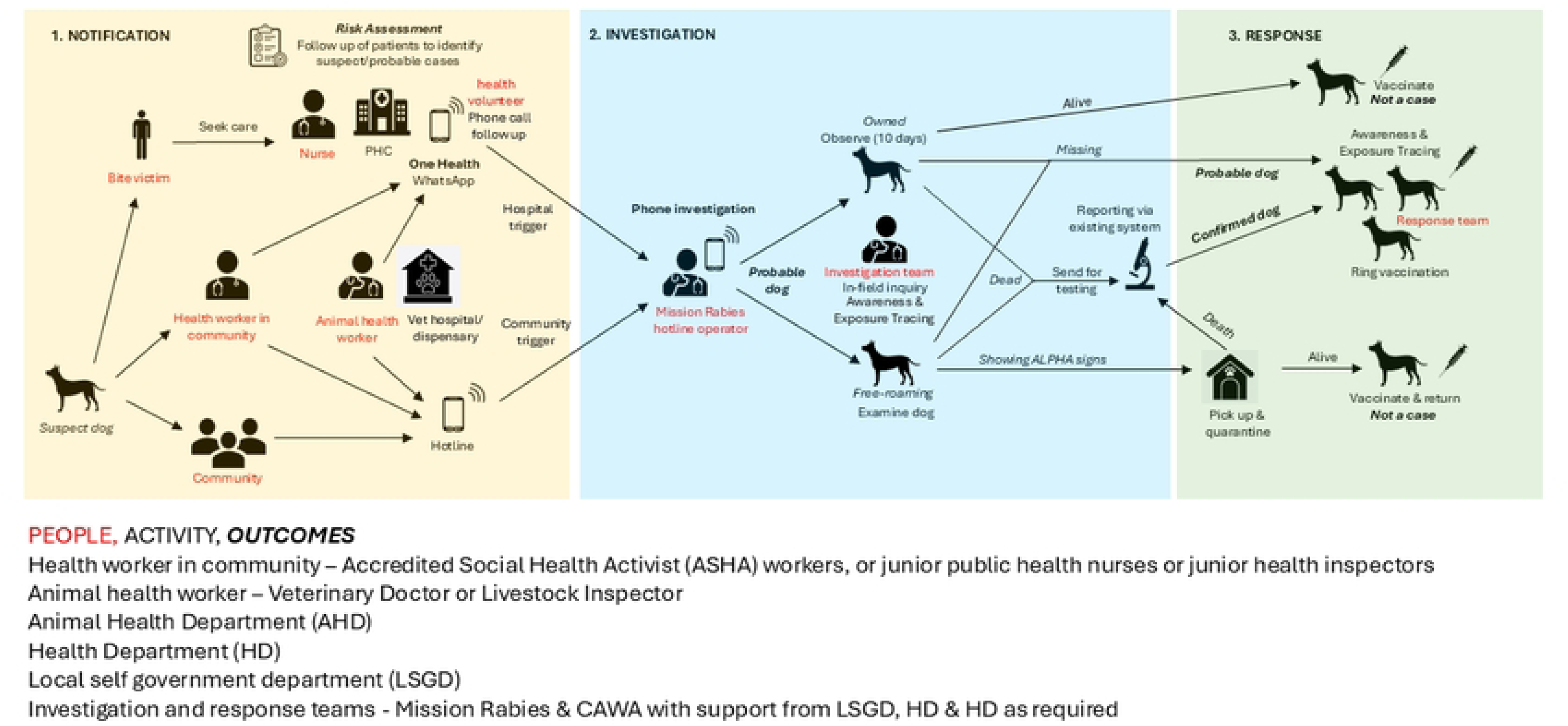
Schematic of IBCM intervention to be implemented for the RCT. Health worker in the community are Accredited Social Health Activist (ASHA) workers, or junior public health nurses or junior health inspector; Animal health workers are Veterinary Doctors or Livestock Inspectors. PHC are Primary Health Centres. Investigation and response teams comprise Mission Rabies & CAWA staff with support from the Local Self Government Department (LSGD), Health department (HD) and Animal Health Department (AHD) as required.

### The IBCM Intervention

*Overview:* We developed an IBCM protocol based on WHO’s Open Course Module 8 on IBCM[12] making adaptations for implementation in Kerala, so that the IBCM risk assessment is not used for PEP decision-making. The IBCM intervention comprises *notification*, *investigation* and *response* steps that formally link the public health and animal health stakeholders, including implementing partners from the NGOs Mission Rabies and CAWA (Fig 3). Notification is designed to occur via two routes, via health facilities through a coordinated One Health WhatsApp group involving stakeholders from health, animal and NGO sectors or via a free public Mission Rabies hotline for veterinary or health personnel or community members to report suspect rabid animals. The WhatsApp notification will follow a risk assessment to assess whether the biting animal is suspicious for rabies (i.e. beyond the national protocol for assessing the wound and PEP needs). The investigation step is designed to determine the animal’s rabies status including laboratory testing and identification of other in-contact individuals requiring PEP. The response step involves collecting and quarantining free-roaming dogs showing rabies signs, and ring vaccination around probable/ confirmed dog rabies cases, as well as awareness raising in affected communities.

We will use the following case definitions[12,23]: a *suspect* rabid animal must show at least one ALPHA sign (A- Aggression, L- lethargy, P- Paralysis, H- Hypersalivation and A-Abnormal Vocalisation) at the time of or within 10 days of the exposure; a *probable* rabid animal is a suspect animal with additional history of a bite by another suspect/ probable animal and/ or is killed/ died/ disappears within 4-5 days of showing signs. A *confirmed* rabid animal is a rabies suspect/ probable animal that is confirmed in the laboratory. An animal is defined as not a case if it survives a 10-day observation, or is a suspect/ probable case ruled out by laboratory testing.

The intervention will engage representatives from the Directorate of Health Services, Directorate of Medical Education, Animal Husbandry Department, local self-government bodies, community health workers, and animal health stakeholders Mission Rabies and Compassion for Animal Welfare Association (CAWA), ensuring a multi-sectoral approach aligned with One Health principles.

#### Intervention components

##### Notification

The notification step involves triggers for investigations from a PHC (hospital trigger), whereby a suspected, probable, or confirmed rabies exposure reports to the facility, or from the community (community trigger) when a dog bites more than one person or animal, i.e. a cluster of bites, or any dog is reported as probable or confirmed rabies, via the hotline (Fig 3). For the community trigger, any member of the community or animal health professionals can report via the hotline.

At the PHC patients will first be assessed by the designated medical officer to determine their PEP requirements, based on the 2019 National Guidelines[24] and a nurse will administer PEP according to the standard of care. The nurse will then undertake an IBCM risk assessment by asking the bite patient a series of questions about the biting dog adapted from WHO guidance.[25] Volunteer health workers will subsequently call these patients over the phone on days zero, 10 and 28 to encourage completion of the intradermal rabies vaccination (IDRV) schedule and follow up on the health and vaccination status of the dog.

The day zero call will be used to identify additional individuals bitten by the same dog and the bite victim will be requested to encourage these others to seek care. If there are multiple bites in the community by the suspect/ probable dog these events will be passed on to the response team via the One Health WhatsApp group (hospital trigger, Fig 3). During the call patients will be educated on ALPHA signs to identify rabies and be asked whether the dog is alive and observable, or has died, been killed or is missing. For dogs that are observable, patients will be requested to monitor the dog at home for 10 days and immediately report via the hotline if any ALPHA signs appear. If the dog is suspect or probable for rabies, meaning the dog has died, been killed or disappeared, or ALPHA signs are reported during the observation period, a response will be initiated. The day 10 follow-up call will check if the dog remains alive and if it is unvaccinated, owners will be urged to vaccinate the dog at the nearest veterinary hospital. The day 28 follow-up call will determine whether the patient has completed their PEP schedule and, if applicable, whether dog vaccination was initiated.

##### Investigation

Following a hospital or community trigger, the hotline operator will begin by calling to clarify the dog’s health and confirm whether and how an in-field inquiry should take place. If the call confirms the need for an in-field inquiry, a response team will travel to the site and examine the dog. If it is already dead the animal will be sent for laboratory testing. If the animal is alive but owned, the owner will be requested to observe the animal under home quarantine to ensure it remains healthy, or call the response team if it shows ALPHA signs or dies. During the investigation community sensitization will be undertaken, including distribution of awareness materials and any other in-contact individuals will be identified and counselled.

##### Response

For any dog identified as probable or confirmed as rabid, a response will be undertaken. Awareness sessions will be done door-to-door and in group sessions within communities including distribution of materials (pamphlets and hotline cards). Ring vaccination of all dogs in the radius of one kilometre around the site of the identified rabid dog will be undertaken with the guidance of the Worldwide Veterinary Services (WVS) app.[26,27] If the dog is alive and free-roaming but showing ALPHA signs, the response team will pick up the dog to take it to the isolation facility for monitoring. If the dog dies it will be sent to the laboratory for testing. If the dog remains alive and healthy for ten days, it will be vaccinated and returned to the community.

### IBCM implementation

Detailed roles and responsibilities of stakeholders implementing IBCM are outlined in Table 2. A One Health WhatsApp group will be set up for each cluster at the start of the study, enrolling relevant stakeholders from PHCs and the Local Self Government Departments (LSGDs). Throughout the study the enrolled nurse at each PHC will enter minimal trigger details for the two patients recruited daily into the One Health WhatsApp group, while all other information will be entered in the ODK form.

**Table 2:** Responsibilities of stakeholders.

| Stakeholder | Responsibilities |
| --- | --- |
| Directorate of Health Services<br>State and District Program<br>officers | Leaders of the program. All communications to the primary care level (PHC/FHC) will be channelised through them |
| Medical officers | Implementation at the primary health care level<br>Wound categorisation Risk assessment<br>Coordination with animal health personnel |
| Health workers, ASHA | Community education, monitoring and reporting of events in households, work in liaison with project field staff |
| Project field staff | Documentation, data entry, monitoring & surveillance, linkage between the human and animal health personnel |
| Directorate of Animal<br>Husbandry | Leaders of the program. All communications to the primary care level (government veterinary hospital medical officer and team) will be channelised through them |
| State Institute of Animal<br>Diseases | Investigations – Lab tests antibody titres post vaccination and testing of rabies in suspect animals, quality control of sample collection, storage, transportation and testing, timely reporting of results |
| Indian Veterinary Association,<br>Kerala | Mobilising support at the field level from veterinary personnel, advocacy and training |
| Government Veterinary<br>Hospital Medical Officers | Assessment and monitoring of the health status of the dog- initial and when required, Communication with the PHC Medical Officer, Supportive supervision of the field staff and livestock inspectors |
| Livestock inspectors & trained<br>dog catchers |  |
| Global collaborator from the<br>University of Glasgow | Technical Consultancy in implementation, training, Cost-Effectiveness analysis |
| Mission Rabies & Compassion<br>for Animal Welfare<br>Association (CAWA) | Technical Consultancy in implementation, training in IBCM, Animal Surveillance and Response, Pick up, Assessment and monitoring of health status of Free-roaming dog during quarantine/observation, sample collection for testing suspect free-roaming dog, Ring Vaccination around probable and confirmed rabies dogs |
| Local Self Government -<br>Political leaders, Officials in<br>the block panchayat | Community engagement, advocacy, ensuring sustainability, social audit |
| Co-investigator from Achutha<br>Menon Centre for Health<br>Science Studies, Kerala | Support with the Open Data Kit data entry form development, storage and maintenance of data |
| Principal Investigator and<br>Team at School of Public<br>Health, Kerala University of<br>Health Science, Department of<br>Medical Education-Government Medical College,<br>Kollam | Overall planning, coordination and management of the project (technical & financial), submission of reports periodically, documentation and convening the meetings with stakeholders.<br>Conduct of workshops, Development of software for data entry, Engaging students posted in the community trainings, Dissemination workshops and Policy briefs, Incorporation into curriculum of medical and paramedical students |

At the start of the intervention in each cluster, relevant stakeholders including both health and veterinary personnel, will undergo joint training on IBCM, followed by ongoing support for implementation, including provisions for surveillance tools, testing, and reporting systems. A package of awareness materials will be distributed to reinforce messages on dog bite prevention, rabies awareness, and the importance of vaccination of both bite victims and dogs. At the trainings, all participants (staff from the enrolled PHCs, veterinary hospitals or dispensaries, and attending community members such as ASHAs and community leaders etc) will be requested to add the hotline number to their phone and will be oriented to report community trigger events to the hotline number (Fig 3). A community awareness programme will also implemented in each cluster at the onset of the intervention. This includes sensitization of key stakeholders such as LSGD representatives, and repeat sessions will be conducted as necessary.

During the sustenance phase, the implementation of IBCM will be monitored across all clusters. Ongoing support and technical guidance will be provided as needed to ensure continuity and fidelity to the intervention.

### Outcomes

The study is guided by the RE-AIM implementation framework (Table 3). The primary outcome *Effectiveness* will be measured quantitatively, through changes in PEP uptake, surveillance and dog vaccination indicators across intervention and control cluster periods, and qualitatively through stakeholder perspectives. The primary health outcome will be bite patient completion rates for PEP, and the primary surveillance outcome will be dogs tested for and confirmed rabid (numbers and percentage change). The secondary health outcomes will be unvaccinated owned biting dogs that were vaccinated (percentage) and numbers of dogs vaccinated during ring vaccination around probable or confirmed cases.

**Table 3:** RE-AIM Framework.

|  |
| --- |
| **REACH** |
| Number of trigger events in the community during the period/ Number of events in which the model could be implemented |
| Number of |
| Qualitative methods to understand reach and/or recruitment – IDI and FGD |
| **EFFECTIVENESS** |
| % change in patients completing PEP, % change in the number of dogs seen by veterinarians and quarantined, % change in the number of dogs vaccinated, % change in the suspect animals tested. These will be compared in the intervention and control cluster periods. |
| Perceived change in scores of Animal welfare at the group level from baseline after the IBCM implementation |
| Measure of robustness across subgroups – comparison of the rural, urban, and tribal clusters |
| Qualitative assessment of effectiveness from stakeholders– IDI and FGD |
| **ADOPTION- SETTING LEVEL** |
| Setting Exclusions (% or reasons) |
| The percentage of settings that participate out of those approached |
| **ADOPTION-STAFF LEVEL** |
| Staff Exclusions (% or reasons) |
| Percentage of staff invited that participate |
| % of doctors who followed the IBCM components among those trained to deliver the service |
| Qualitative assessment of adoption from stakeholders– IDI and FGD |
| **IMPLEMENTATION** |
| Percentage of “perfect” IBCM components implemented |
| Adaptations made to IBCM during study |
| Cost of IBCM (time or money) |
| Consistency of implementation across staff/time/settings/subgroups (not about differential outcomes, but a process) |
| Use of qualitative methods to understand implementation- IDI and FGD |
| **MAINTENANCE- INDIVIDUAL LEVEL** |
| Calls received by the mission rabies /central coordinating office three months after the completion of implementation as compared to the period in which the intervention was ongoing |
| **MAINTENANCE-SETTING LEVEL** |
| % change in the number of dogs vaccinated, % change in the number of suspect animals tested – cluster level data after three months of completion of the intervention |

The secondary surveillance outcome will be the change in dogs examined by a veterinarian, quarantined, and subsequently vaccinated once determined to be healthy (numbers and percentage). Numbers of additional bite victims identified through the phone call follow up will be examined as an exploratory outcome. Potential for cost savings will be measured through the percentage change in patients who would not require PEP, or could be administered PEP at reduced cost (no RIG, or no final dose) had judicious PEP administration protocols been implemented. These outcome indicators will be compared before and after IBCM implementation in each cluster.

*Reach* will be examined through the coverage of trigger events. *Adoption* will be assessed at the setting and staff levels, capturing participation rates, characteristics of adopters, and fidelity to IBCM components. As part of adoption, client satisfaction will be assessed using structured questions integrated into the evaluation tool. *Implementation* will be assessed based on the degree of adherence, adaptations, cost, and consistency across contexts, supplemented by in-depth interviews and focus group discussions. Finally, *Maintenance* will be evaluated at both the individual and setting levels through follow-up of patient behaviours and health system performance indicators beyond the intervention period over the two-month follow-up period post-intervention, focusing on continued delivery of services, adherence to the implementation protocol, and sustained individual behaviour change. These indicators will be monitored through weekly Integrated Disease Surveillance Program (IDSP) review meetings, which are regularly conducted with doctors and healthcare workers at each PHC.

### Data Collection and Analysis

Risk assessment data for the two bite patients recruited daily to the study at each enrolled PHC will be recorded by nurses. Data on each of the follow up calls (day 0, 10 and 28) to bite patients will be collected by the health volunteer. The risk assessment (S2) and phone call follow up (S 3) collection tools will be converted into electronic forms using the Open Data Kit (ODK) platform. Data on the investigation and response will be collected by the investigation and response teams in the WVS app.

Comparisons between intervention and control periods will be conducted using Generalized Linear Mixed-Effects Models that account for intra-cluster correlation for each of the trial outcomes (Table 3). Subgroup analysis will be done across clusters and urban-rural settings. We hypothesize that PEP completion rates of recruited patients will increase under IBCM, and that confirmation and vaccination of otherwise unvaccinated healthy dogs will increase, as will detection of probable rabid dogs and laboratory confirmation of cases. We expect that rapid responses and associated ring vaccination following case detection will also increase with IBCM. Data from the risk assessment will be used to calculate the potential for cost savings through judicious PEP. Answers to the risk assessment questions will be scored and computed to identify whether the individual required PEP as per the WHO risk assessment. The proportion of PEP that could have been avoided will be used in the cost-effectiveness analysis.

The costing will include both direct costs, such as PEP, diagnostics, and logistics, and indirect costs, including productivity losses and time costs. Data will be obtained from government records, hospital datasets, stock inventories, and expert consultations, with annualization applied to reflect typical implementation costs.

Qualitative data collection will involve In-depth Interviews (IDI) with political leaders at the state (1), district (1), and local (6) levels, program managers from both the human and animal health sectors (1 each at state and district levels), medical officers and health care workers from both the DHS and the AHD, ASHA workers and IBCM investigators, as well as individuals exposed to animal bites for beneficiary case studies. Data collection will guided by semi-structured IDI and FGD guides (S4 & S5), developed to capture key implementation outcomes. IDIs and FGDs will be scheduled at the convenience of the participants, with prior appointments and permissions and will be conducted in settings that ensure privacy and comfort for participants. All interviews and FGDs will be recorded with prior consent, with 5% of sessions directly supervised.

Recordings of IDIs and FGDs will be transcribed verbatim and translated into English when required, with 5% of transcripts cross-verified against recordings, with replacements made when necessary. Thematic analysis will be undertaken using both manual methods and NVivo software to ensure systematic organization and rigor. Responses will be grouped to identify domains of homogeneity across different stakeholder categories, facilitating comparative analysis. Coding and categorization will be applied to each domain and representative quotations highlighted for depth. Results will be synthesized into an ethnographic summary that integrates thematic patterns with quotable narratives, to provide nuanced insights into stakeholder experiences and perceptions.

A cost-effectiveness analysis will be conducted to evaluate the Integrated Bite Case Management (IBCM) intervention compared with the status quo. The primary outcomes will include human rabies deaths averted, corresponding disability-adjusted life years (DALYs) averted, animal rabies cases detected, cost per animal rabies case confirmed, and cost savings attributable to judicious use of post-exposure prophylaxis (PEP through application of the WHO rabies risk assessment framework).

Effectiveness inputs will be derived from the stepped-wedge cluster randomised trial and incorporated into a stochastic rabies transmission model linked to a probabilistic decision-tree model to capture human and animal health pathways. Model parameters will be informed by published literature, routine surveillance data, and findings from the primary study.

Two scenarios will be modelled. The status quo scenario will assume provision of PEP without systematic application of the WHO risk assessment chart and continuation of current levels of mass dog vaccination. The One Health IBCM scenario will assume systematic implementation of the WHO risk assessment chart, integration of IBCM between human and animal health sectors, and continuation of mass dog vaccination.

Incremental cost-effectiveness ratios (ICERs) will be calculated and compared against relevant country-specific cost-effectiveness thresholds. Parameter uncertainty will be explored using one-way sensitivity analyses and probabilistic sensitivity analysis employing Monte Carlo simulation.

The base-case analysis will adopt a five-year time horizon (2025–2030) to capture medium-term outcomes of IBCM scale-up, with an additional ten-year horizon modelled to assess longer-term impacts. Both costs and health outcomes will be discounted at an annual rate of 3%. Analyses will be conducted from both the healthcare system and societal perspectives. Costs will be reported in Indian Rupees and converted to US dollars using the exchange rate prevailing at the time of project initiation.

The overall study design is summarised in Table 4

**Table 4.**
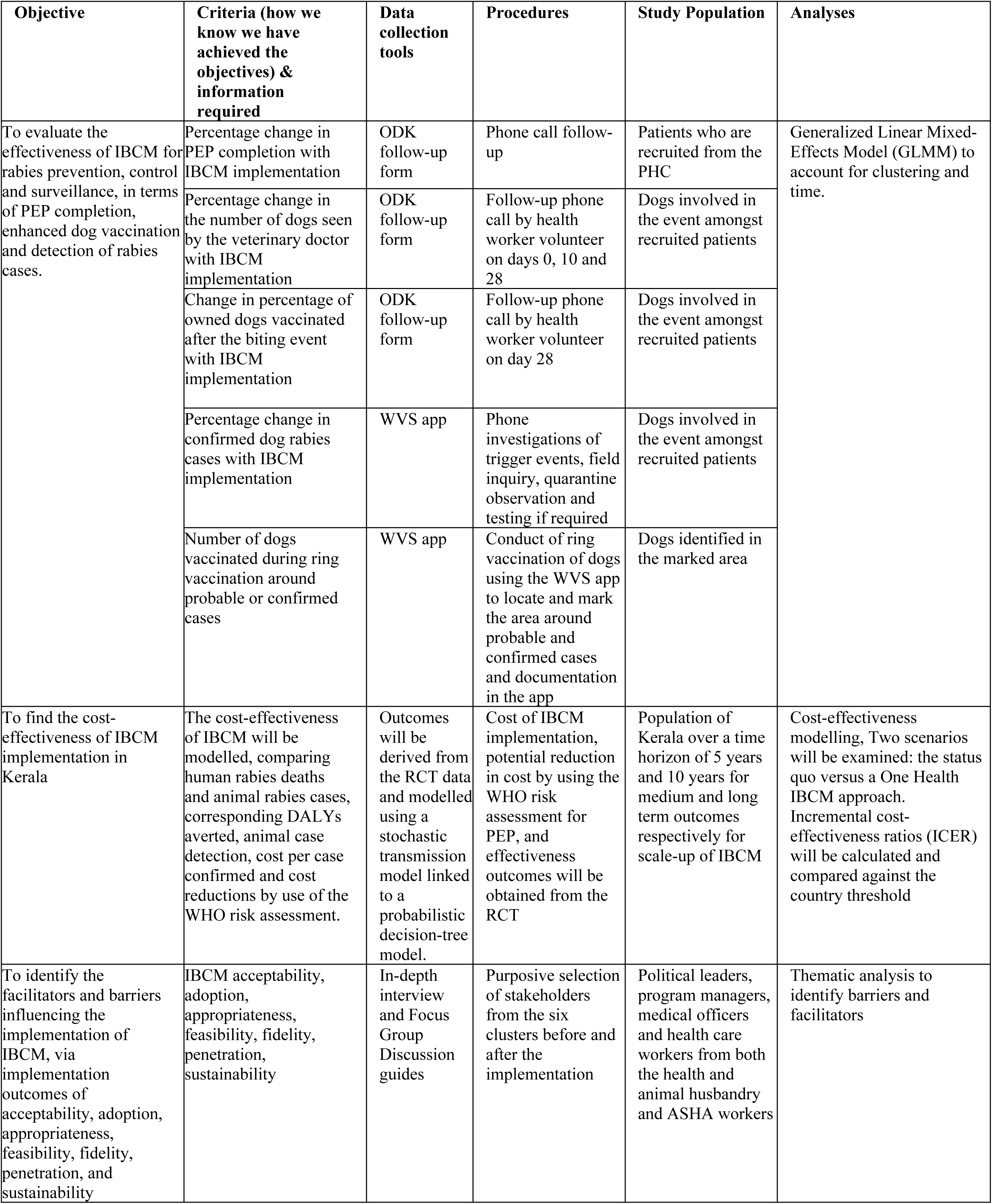
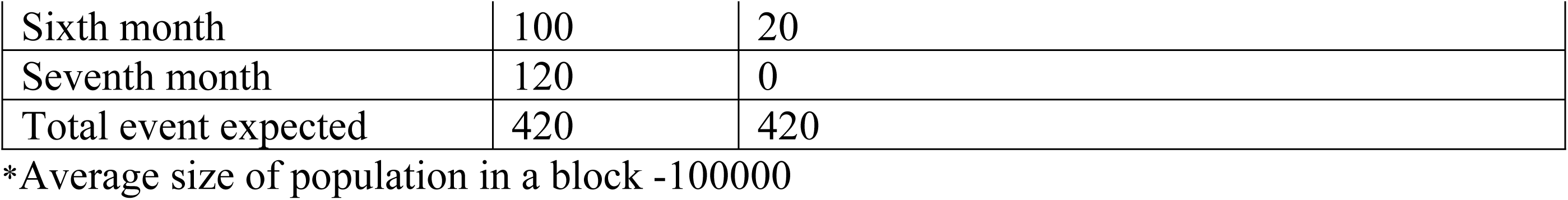
Study design for data collection and analysis to determine whether IBCM achieved the desired outcomes.

### Sample size

The sample size for the baseline cross-sectional survey is based on national (1.96%) and local (Thiruvananthapuram: 1.5%) estimates of the annual incidence of animal bites. Using the formula for a calculating a proportion with 95% confidence (4pq/l2); taking p as 1.5 (p is the annual incidence estimate of animal bites, q-98.5 (100-p), and l (precision) as an absolute 2; the required sample size is 148, which, when adjusted for a design effect of 2, becomes 296. Assuming 15% for non-response, increases the total sample size to approximately 350 individuals. Given an average household size of 4, a total of 88 households will be surveyed. To ensure adequacy, we propose surveying 100 households per cluster and therefore 600 households in the six clusters together.

Given that Category II bites are the most commonly managed at the PHC level, they are used as the basis for estimating the expected number of events (recruited bite patients) during the RCT. The projected number of bite cases is detailed in Table 5.

**Table 5:**
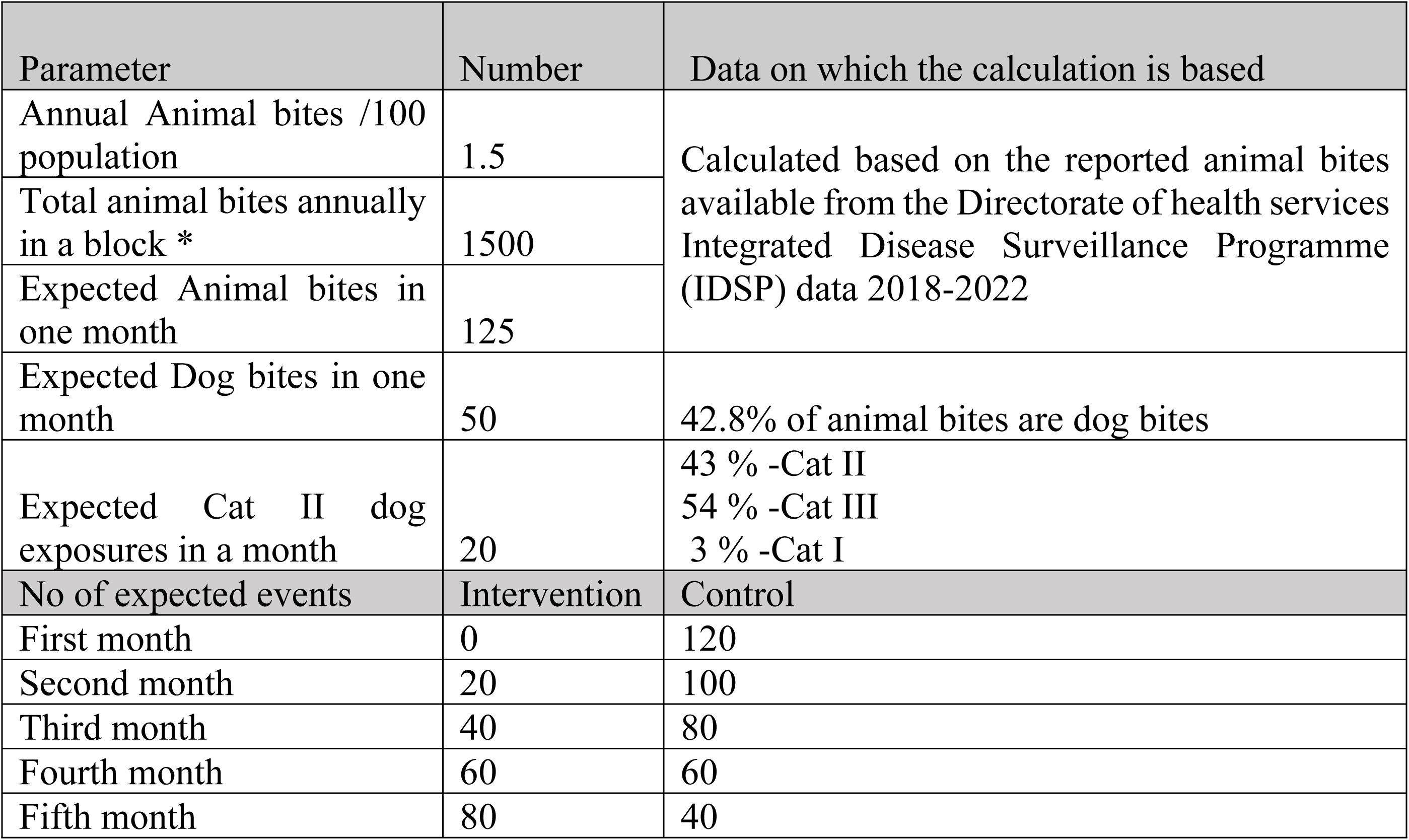
Expected number of events during the implementation period.

We estimated the RCT sample size to be sufficient to detect a 15% decline in potential PEP requirements after IBCM implementation among those seeking care, using Pocock’s formula [22] for comparison of proportions and incorporating a design effect. The expected proportion of events in the control arm was set at 0.75, and in the intervention arm at 0.90, corresponding to a ratio of 0.833.

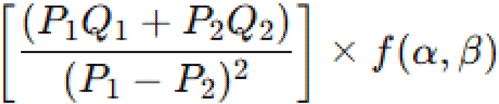

Where: *P*_1_ = 0.90 and is the proportion expected to start PEP prior to IBCM, while *P*_2_ = 0.7 corresponds to a reduced proportion requiring PEP under IBCM. *Q*_1_ = 1 - *P*_1_ = 0.10; *Q*_2_ = 1 - *P*_2_ = 0.25 and *f*(*α,β*) = 16, derived from 2 × 8 for a significance level of *α* = 0.05 and power 1 - *β* = 0.80. We assume a design effect of 1.95, DE = 1+(number in each cluster-1) x ICC (Intra Class Correlation); based on an average cluster size cluster of 20 and an ICC of 0.05. Thus, the required sample size per arm is 385, trigger events which is feasible considering the anticipated number of events (n = 420).

We further estimated the sample size for a stepped-wedge cluster-randomised design using nQuery Advisor (version 8.6). The sample size was calculated to detect a difference in proportions between the control and intervention periods. Assuming six clusters, with each cluster contributing 140 participants over seven time periods (approximately 20 new participants per cluster per period), the study has more than 90% power to detect an absolute increase in the outcome proportion from 0.75 in the control condition to 0.90 in the intervention condition at a two-sided significance level of 5%. An intra-cluster correlation coefficient of 0.05 was assumed. Based on these assumptions, the total required sample size was 840 participants.

For the process evaluation, a stratified purposive sampling technique is used to ensure representation across stakeholder categories and performance levels (e.g., best- and least-performing ASHA workers). A total of 46 in-depth interviews will be conducted across the six study clusters (Table 6). Six FGDs with practising health workers will be conducted before and after IBCM implementation, one in each cluster during both time periods, yielding a total of 12 FGDs. Two beneficiaries per cluster (one before and one after the IBCM implementation), preferably dog owners with bite exposure experience, will be selected for detailed case studies and experience sharing.

**Table 6:** Number of In-depth interviews with stakeholders.

| Sl. No. | Category | Numbers |
| --- | --- | --- |
| 1. | Political Leaders | 8 |
| 2 | Program Managers | 2 |
| 3. | Medical Officers -Health Department | 6 |
| 4. | Medical Officers- Animal Husbandry Department | 6 |
| 5. | Health Care Workers<br>IBCM supervisor Health Department | 6 |
| 6. | Health Care Workers<br>(IBCM supervisor) Animal Husbandry Department | 6 |
| 7. | Accredited Social Health Activist (ASHA) * | 12 |
| Total |  | 46 |

\*ASHA – voluntary workers selected from the community working for the health

### Ethical Approvals

The study is approved by the WHO Ethics Committee (2024.9.Ind), Kerala University of Health Sciences (AX04/SOP07A/V2) and the ethics committee of Government Medical College, Thiruvananthapuram (HEC No.12/06/2024/MCT). Permissions have been obtained from the departments of health (GO(Rt)No. 2287/2024/H&FWD), animal husbandry (DAHO/TVM/409/2025-g) and local self-government (LSGD/PD/6303/2024-PH2). Approval was also obtained from the Government of India (F.No.P-29010/41/2024-DM) for international collaboration.

Informed written consent will be obtained from all participants and stakeholders. Only de-identified data, with no personal identifiers will be used for analysis. Personal information will solely be used to facilitate appropriate follow-up and to ensure the provision of necessary care to patients, families, animals, and communities involved. Animal welfare will be prioritized during the study.

The study protocol has been reviewed and aligned with the StaRI (Standards for Reporting Implementation Studies) guidelines to ensure comprehensive reporting of all relevant components (S6)

### Status and Timeline

The data collection started on 1.2.2025 with the survey to estimate the animal bite frequencies and treatment-seeking. The baseline assessment started on 1.03.2025, intervention on 1.04.2025 and the study is expected to be completed by 30.01.2026, with an extension of the sustenance phase for an extra month. Accordingly ethics committee is informed and an extension for continuation obtained (UEC/5/KUHS/1/2024(version2))

## Discussion

IBCM is increasingly recognized as a valuable One Health surveillance approach, fostering collaboration between human health and veterinary services to ensure timely detection, reporting, and response to suspected rabies exposures. In Kerala, where human–dog interactions are frequent and rabies remains endemic,[28] IBCM can serve as an essential bridge linking bite patient management with animal surveillance. By strengthening veterinary reporting and laboratory confirmation of rabies in animals, the approach can guide immediate public health responses while informing targeted dog vaccination campaigns, thereby addressing the root source of transmission and advancing progress towards the state’s 2030 rabies elimination goals.

A distinctive feature of our protocol is the adaptation of IBCM to the local policy and stakeholder context. While WHO’s IBCM guidance emphasizes rationalized use of PEP by linking administration to animal rabies investigation outcomes, this component is not prioritized in our study. Stakeholder consultations revealed reluctance to reduce PEP administration, given divergence from current national rabies prevention guidelines, which emphasize universal PEP coverage. Accordingly, our IBCM approach focuses on strengthening animal surveillance, ensuring rapid dog vaccination response to confirmed cases, and generating actionable data for targeted interventions. This shift underscores the value of contextual tailoring in implementation research while maintaining the One Health ethos. The IBCM approach evaluated in this study differs from the Goa model, which primarily utilizes public and veterinary reporting through a hotline mechanism. Our model initiates IBCM within the health sector by identifying cases from patients presenting with dog-associated bite injuries, an approach that may allow more comprehensive one health approach. It also enables capture of more events given the higher likelihood of reporting through health facilities

IBCM has been found to greatly increase case detection of rabies across a range of settings. In Tanzania, increased detection of probable rabid dogs was attributed to the collaboration fostered between health workers and veterinary officers.[5] In Albay Province of the Philippines, implementation of IBCM quadrupled rabies case detection compared to passive surveillance.[29]similar to the experiences from Uganda.[30]

In terms of improving best practices for PEP administration, IBCM has both potential to reduce costs through judicious use of PEP and to improve adherence to PEP regimens amongst those exposed to rabies.[31] Critically, through systematic sharing of information across sectors to ensure coordinated action, our implementation study aims to increase both cross-sectoral coordination and community engagement.

The stepped-wedge design is suited to this evaluation, allowing phased introduction across districts so that all eventually benefit while providing comparative data on effectiveness and feasibility. This design is ethically favourable and operationally pragmatic, capturing heterogeneity in local contexts and enabling iterative learning during rollout.[32–34] Implementation research methods allow systematic exploration of facilitators, barriers, and contextual factors influencing integration into existing health and veterinary systems, ensuring that findings are not only about effectiveness but also about scalability and sustainability.[35, 36]

We anticipate challenges, including under-reporting of animal rabies cases, variable intersectoral coordination, and logistical delays in laboratory confirmation. Community-level concerns, including trust in veterinary reporting and responsiveness of rapid dog vaccination teams, may also influence outcomes. These limitations will be documented to inform adaptation and scaling. Dissemination will occur through policy briefs, stakeholder workshops, and peer-reviewed publications, with special emphasis on engaging state-level One Health platforms to translate findings into policy and programmatic action. Amendments to the study protocol, including potential termination, will be reviewed and approved by the institutional ethics committee, with due regard for participant safety, transparency, and reporting requirements.

## Data Availability

This is a protocol and no results are presented based on any data analysis

## Supporting Information

S1**-** This is the data collection form in ODK used for the cross-sectional survey

S2- This is the data collection form in ODK used by the nurses to get the details of the recruited events and doing the WHO risk assessment for PEP

S3- This is the data collection form in ODK used by the health volunteers for follow up of the patients and the dogs

S4- This file contains the IDI schedules used for the In-depth interviews

S5- This is the FGD guide

S6- This is the STARI checklist showing the page numbers in which the items are described

## Data availability

No datasets were generated or analysed for this protocol. Data arising from the study will be made available upon reasonable request from the corresponding author, subject to institutional and ethical approvals, after de-identification.

## Acknowledgements

We gratefully acknowledge Dr. Mohanan Kunnammal, Vice Chancellor, Kerala University of Health Sciences (KUHS); The authors wish to express their sincere gratitude to Dr. Rajan N. Khobragade, Additional Chief Secretary, Health & Family Welfare Department, Government of Kerala; Dr. Reena K.J., Director of Health Services, Kerala; and Dr. Reeta K.P., Additional Director of Health Services (Public Health), for their leadership, guidance, and support that enabled this work.

We place on record our gratitude to Dr. C. P. Vijayan, Pro Vice Chancellor; Prof. (Dr.) Gopakumar S., Registrar; Dr. Shaji K.S., Research Dean; and Dr. Binoj R., Academic Dean, KUHS, for their institutional support and facilitation of academic collaboration. We also thank Dr. Suma T.K., ICMR Emeritus Scientist, School of Public Health, for her valuable insights and mentorship.Our sincere thanks are extended to Mrs. Jisharaj V.R., Assistant Professor, School of Public Health, and to Mrs. Nimmy, Mrs. Riya, Mr. Hareesh, and Mr. Athul from the Office of the School of Public Health for their administrative and logistical support.

We thank Dr.Althaf A, Dr.Sunitha, Dr. Anil, Dr. Rameez, Dr. Niju, Dr. Manju, Dr.Latha, Dr. Reshmi, Medical Officers and all the staff and ASHA workers of the health centres where the study is being done.

We also thank Smt. Namitha, Smt. Rajasree from Mission Rabies and Smt, Akshara, Shri. Lionelle from CAWA for the tremendous support throughout.

We are deeply thankful to Dr. Katrin Bote, Technical Officer – Neglected Tropical Disease Control, Department of Communicable Diseases, WHO Regional Office for South-East Asia (SEARO), for her constant technical support and encouragement. We also acknowledge with appreciation the support extended by the entire WHO-SEARO and TDR teams, whose continued engagement greatly strengthened the study.

Finally, we acknowledge the support and collaboration from the team at the School of Biodiversity, One Health & Veterinary Medicine, University of Glasgow, which contributed significantly to the interdisciplinary perspective of this work. We specially appreciate and acknowledge Smitha Abraham, International Project Coordinator, for her coordination and project management support.

